# Difference of brain cortical thickness and area of different lobes between fetuses with intrauterine growth restriction and control group assessed by 3 Tesla MRI

**DOI:** 10.1101/2020.10.31.20222737

**Authors:** Behnaz Moradi, Mahboobeh Shirazi, Zohreh Alibeigi nezhad, Nazanin Seyed Saadat, Hassan Hashemi, Mohammad Ali Kazemi, Masoumeh Gity, Maryam Rahmani, Hossein Ghanaati

## Abstract

**Background:** Intrauterine growth restricted (IUGR) is a major factor of perinatal and long-term morbidity and is associated with abnormal fetal brain development but its pattern of brain involvement is still unknown.

**Materials and Methods:** 42 women with IUGR pregnancy and 28 women with normally-grown fetuses between 28-38 weeks underwent 3Tesla MRI. Cortical thickness was assessed in 4 regions and was corrected by biparietal diameter/2. Also, whole brain surface area (WBA) and areas of 6 brain regions were included and corrected by WBA.

**Results:** IUGR fetuses had significantly thinner cortical thickness in the insula and temporal lobes compared to the control group (0.034 vs 0.043 and 0.036 vs 0.047 respectively, P value of < 0.05). They had significantly reduced WBA (P value: 0.028). The corrected brain areas were not significantly different between groups except for the corrected areas of cerebellum and hippocampus which were increased in IUGR fetuses compared to the control group (0.147vs 0.130,0.017vs 0.0125 respectively, P value of < 0.05).

**Conclusion:** IUGR fetuses had significantly thinner insular and temporal lobe cortex and smaller WBA compared to the control group. Among different brain regions, cerebellum and hippocampus were less affected by growth restriction in utero period.

## Background

Intrauterine growth restriction (IUGR) occurs in 5-10% of all pregnancies (1). IUGR is defined as a birth weight below the 10th percentile for gestational age and is mostly caused by placental insufficiency (2, 3).

Multiple pathophysiologic mechanisms occur during IUGR due to placental dysfunction (4). Following placental insufficiency, oxygen and nutrients transfer to the fetus is reduced, blood flow selectively redistributes to the vital organs including the brain, in contrary, blood flow supply other fetal organs is decreased (2). These hemodynamic changes cause recurrent hypoxemia episodes (4). Evidence shows an adverse effect of hypoxia on brain development along with cell number and size reduction and consequently whole brain weight loss (5). Ultrasound with Doppler evaluation are used in high risk pregnancies for detection of the severity of hypoxia (6).

IUGR is a major factor of perinatal and long-term morbidity and is associated with abnormal fetal brain development, cerebral palsy, mental retardation, neurological dysfunction in motor skill and cognitive disorder (7). These neurodevelopmental dysfunctions are correlated with certain neurostructural changes in the brain (8). These developmental changes are seen in both severe and mild IUGR (9).

There is a major challenge in the perinatal diagnosis of brain injury in IUGR pregnancies (10). MRI has recently opened up the possibility of determining the structural and morphometric differences between the brain structure of IUGR and appropriate growth for age (AGA) fetuses (1,2) Regarding previous researches, the pattern of brain abnormalities associated with IUGR is still unknown (11,12).

Cortical development is a complex process that is affected by many factors (9). MRI in IUGR fetuses shows significant differences in brain volume and cortical characteristics such as gyrification pattern and cortical thickness (8,13,14). Some studies found brain grey matter volume reduction especially in temporal and insular lobes (15).

There are few studies that measure brain cortical thickness and areas in IUGR fetuses. So, the aim of this study was to evaluate the effect of growth restriction on cortical thickness and areas of the fetal brain in different regions compared to controls by 3T MRI.

## Materials and Methods

### Patient Population

We prospectively studied a sample of 42 women with IUGR pregnancy and 28 normal fetuses in the control group with gestational age between 28-38 weeks. This study was performed in the imaging department of our hospitals between March 2017 and January 2019. All participants were given written informed consent and the study was approved by institutional board members.

Gestational age was measured based on the first-trimester crown-rump length in all fetuses. We defined our IUGR cases as fetal weight less than 10th percentile for gestational age by ultrasound and according to the reference data (16).

The control group was chosen from fetuses with fetal and postnatal weight ≥ 10 percentile with similar gestational age to the IUGR group who referred for MRI of extra–CNS pathologies. Maternal demographic data and perinatal data including gestational biometry and weight at the time of MRI session, birth weight and pregnancy outcome after birth were recorded in study groups. MRI

Exclusion criteria were the presence of congenital infection, multiple pregnancies, chromosomal abnormalities or suspicious genetic syndromes, chronic maternal disease, general contraindications for MRI and any structural or brain abnormalities that were detected on MRI. Because of fetal calvarium shaowing, ultrasound is not suitable modality for evaluation of brain lobe areas. On the other hand, ultrasound has not enough soft tissue resolution for differentiating cortex from white metter. Therefore, in this study we assess the brain charactristics of fetuses by fetal MRI. According to ACR guideline, exposure to the 3T magnetic fields used in the routine clinical MRI process has not any reproducible harmful effects on the developing fetus. Also by using rapid acquisition sequences and decrease the acquisition time we tried to decrease fetal exposure (17).

### Doppler Ultrasound

Ultrasound examination was carried out using a transabdominal 2-6 MHz curved-array transducer (Affiniti 50, General imaging configuration, Philips ultrasound machine, USA) during the same week of MRI session.

Fetal umbilical artery (UA) and middle cerebral artery (MCA) pulsatility index (PI) were assessed and values > 95^th^ centile and < 5th centile considered abnormal respectively according to the reference chart (18). Abnormal cerebroplacental ratio (CPR) was defined as a value below the fifth centile. The mean of bilateral uterine artery (UtA) PI was measured by transabdominal sonography and values were considered abnormal when was greater than the 95th centile (19).

IUGR cases were subcategorized into two study groups due to IUGR severity: Group A: with severity sign as fetal weight less than the third percentile and/or abnormal UA or UtA PI and/or abnormal CPR. Group B: without above mentioned criteria.

### MRI protocol

Fetal brain MRI was carried out at a 3T MR unit General Electric system (GE Healthcare, discovery 750 GEM), using a 16-channel phased-array coil. Brain MRI was performed within the same week of diagnosis of IUGR by ultrasound.

Imaging process performed after 4 hours of fasting and without sedation, while the patient was positioned in left lateral decubitus. The duration of brain MRI was around 20-30 minutes per case. The examination protocol applied to study patients included the single shot fast spin echo T2-weighted sequences with slice thickness 4mm with no gap, field of view 25-27cm, repetition time 1500 milliseconds, echo time 98 milliseconds, matrix 288 X192 mm, flip angle 180 degrees, and acquisition time 25 seconds. Brain MRI was taken at 3 different orthogonal planes. If motion artifact degradation had been detected, the sequence was repeated until a satisfactory image was obtained.

### MRI analysis

Offline analysis of all morphologic and biometric brain measurements was performed on Infinite Picture Archiving and Communication System (PACS) by an experienced specialist. The radiologist was aware of the preliminary diagnosis of IUGR but not the subgroups. Brain biometry [biparietal diameter (BPD) and head circumference (HC)] were measured in the transthalamic axial plane and were assessed based on standard reference chart (16).

### Brain cortical thickness assessment

Insular cortex thickness: in the axial plane just below the plane of cavum septum pellucidum; Frontal cortex thickness and occipital cortex thickness: in the axial plane at the level of cavum septum pellucidum; Temporal cortex thickness: In the coronal plane, just anterior slice to pons level. Two parts of it was measured from inner to outer. We calculated the average of bilateral cortical thickness for each region, then we corrected them by dividing by BPD/2 (Figure. 1).

**Figure. 1.**
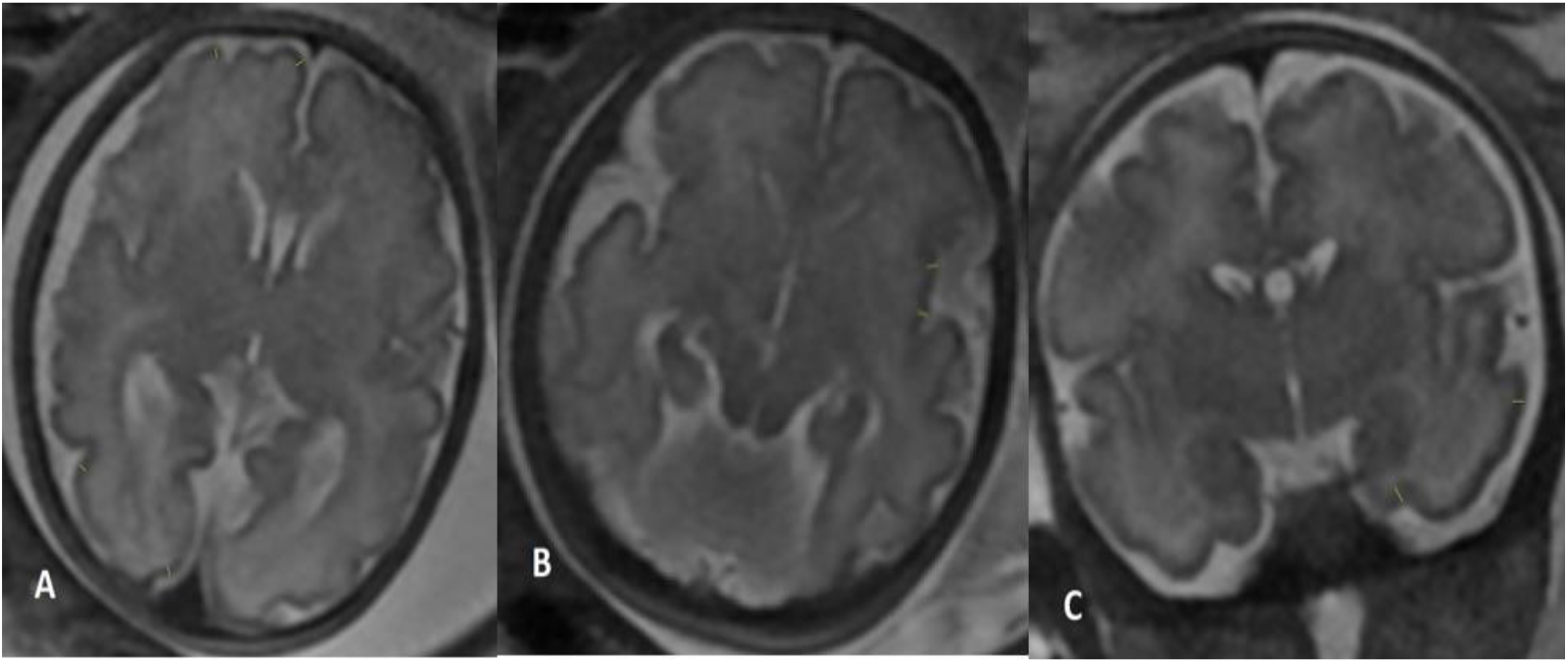
T2-Weighted magnetic resonance images show brain cortical thickness assessment. (a) Frontal and occipital lobes cortical thickness measurement (in the axial plane at the level of cavum septum pellucidum), (b) Insular cortical thickness measurement (in the axial plane just below the plane of cavum septum pellucidum), (c)Temporal lobe cortical thickness measurement (In the coronal plane, just anterior slice to pons level). Two parts of them were measured from inner to outer

### Brain area assessment

Occipital lobe area: in the first para-sagittal plane, near falx, posterior to the parieto-occipital sulcus; Temporal lobe area: in the coronal plane, just anterior to the pons level; Frontal lobe area: in the axial plane, just superior to cavum septum pellucidum and lateral ventricles, anterior to the Rolandic fissure; Parietal lobe area: in the axial plane, immediately superior to cavum septum pellucidum and lateral ventricles, posterior to the Rolandic fissure; Cerebellum: in the axial plane, at the level of the middle cerebellar peduncle; Pons: at the same level described for the cerebellum.; Midbrain: in the axial plane, at the level of the midbrain; Whole brain surface area (WBA): in the axial plane at the level of cavum septum pellucidum; Whole intracranial area: at the same level of WBA by tracing cursor inside the bony calvarium (Figure 2). A lot of effort was put into taking standard and motionless images and oblique images were omitted.

**Figure. 2.**
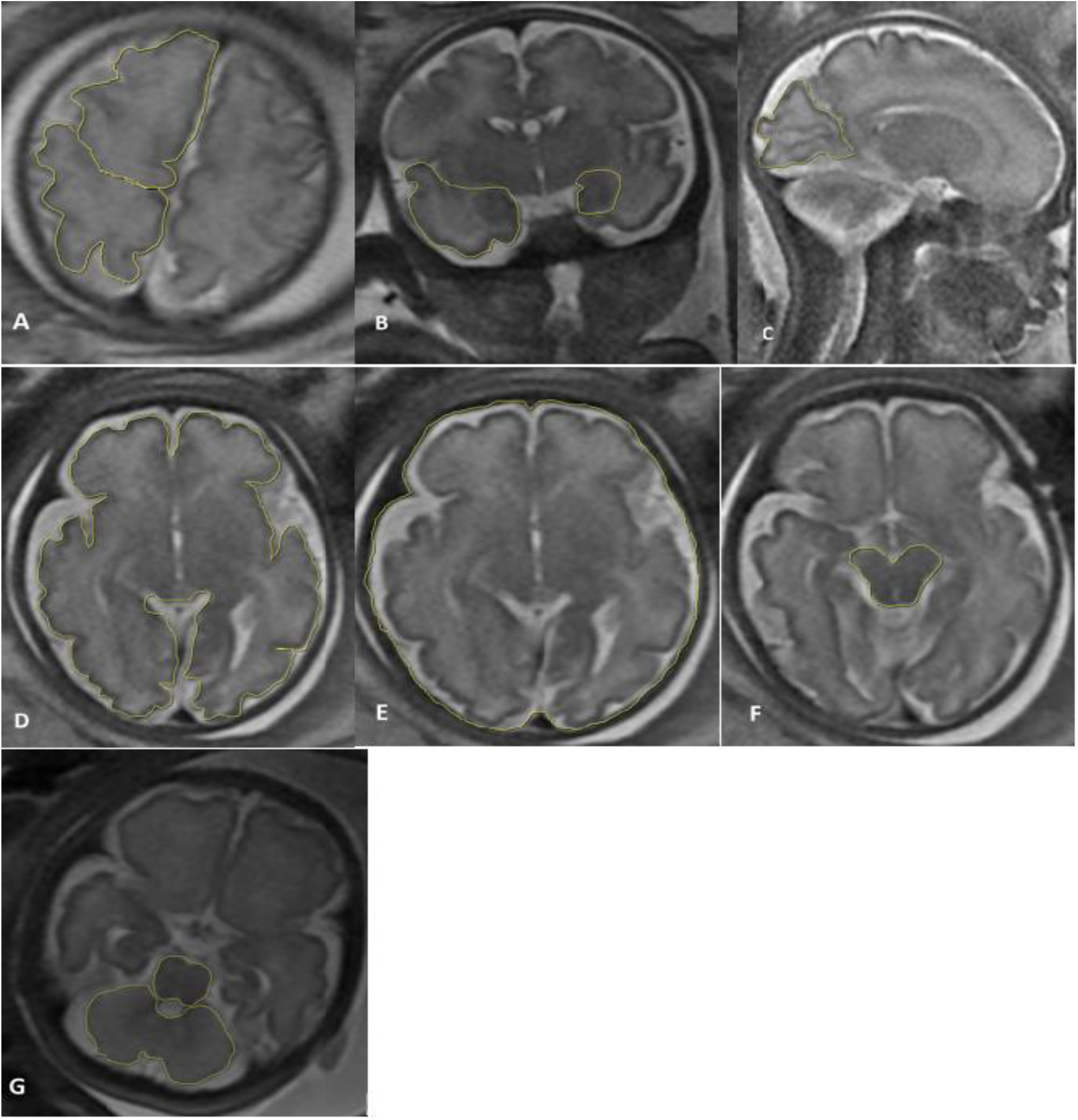
T2-Weighted magnetic resonance images demonstrate different brain lobe area assessments. (a) Frontal and parietal lobe areas (in the axial plane, just superior to cavum septum pellucidum and lateral ventricles), (b) Temporal lobe area (in the coronal plane, just anterior to the pons level), (c) Occipital lobe area (in the first para-sagittal plane, near falx, posterior to the parieto-occipital sulcus), (d) Whole brain area (in the axial plane at the level of cavum septum pellucidum), (e) Whole intracranial area (at the same level of WBA by tracing cursor inside the bony calvarium), (f) Midbrain area, (g) Cerebellum and pons areas (in the axial plane, at the level of the middle cerebellar peduncle).

All measurements were delineated by cursor-guided free-hand trace and performed twice in which the mean of each one was used for analysis. Afterward, the mean of the bilateral area for each region was calculated. The corrected brain area was obtained by dividing each brain area by WBA and corrected WBA was calculated by dividing by whole intracranial area.

### Data analysis

The software package SPSS 22 (IBM Corp., Armonk, NY, USA) was used for the statistical analyses. Data were summarized using means and SDs for quantitative variables and using frequencies and percentages for qualitative ones. In the data analysis, first, the normal distribution of quantitative data was investigated using the Kolmogorov-Smirnov test (K-S test). A comparison between groups was done by using Fisher, Chi-square and T-tests. Values less than 0.05 were considered statistically significant.

## Results

### Demographics and clinical measures

We included a total of 42 fetuses with IUGR and 28 fetuses in the control group with similar gestational age (around 28-38 weeks) in this study. IUGR fetuses consisted of 24 fetuses in subtype A and 18 cases in subtype B. Maternal and fetal characteristics and perinatal outcome of the study groups are presented in table. 1. Only one death (stillbirth) in subtype A was reported. UA PI>95th centile, MCA PI < 5th centile and UtA PI > 95th centile were detected in 11 (45%) fetuses, 4 (16.6%) fetuses and 5 (21%) cases with subtype A IUGR respectively. Absent or reversed diastolic flow in an umbilical artery or abnormal flow in ductus venosus were not found in any of IUGR subgroups (no cases with severe hypoxia).

**Table 1:**
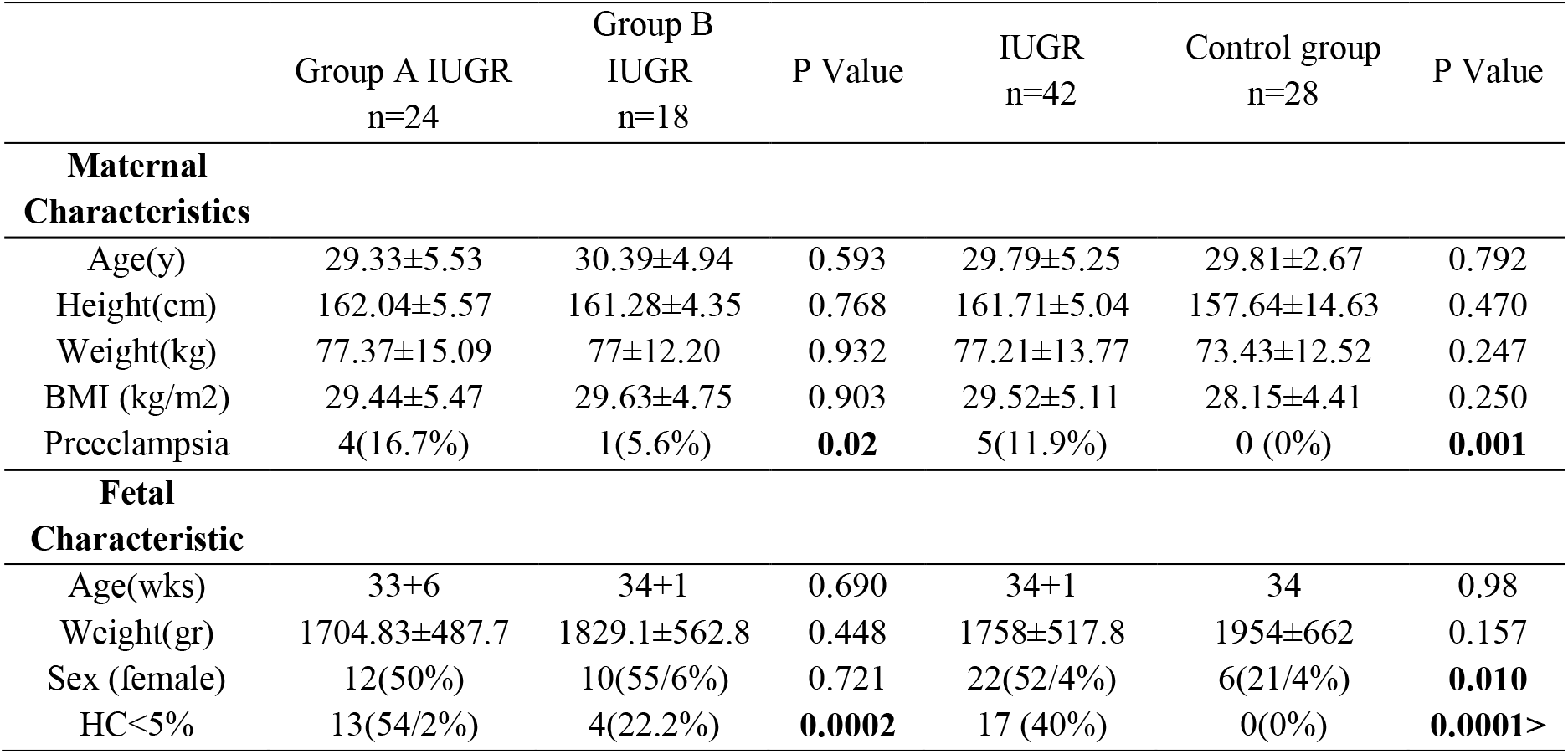
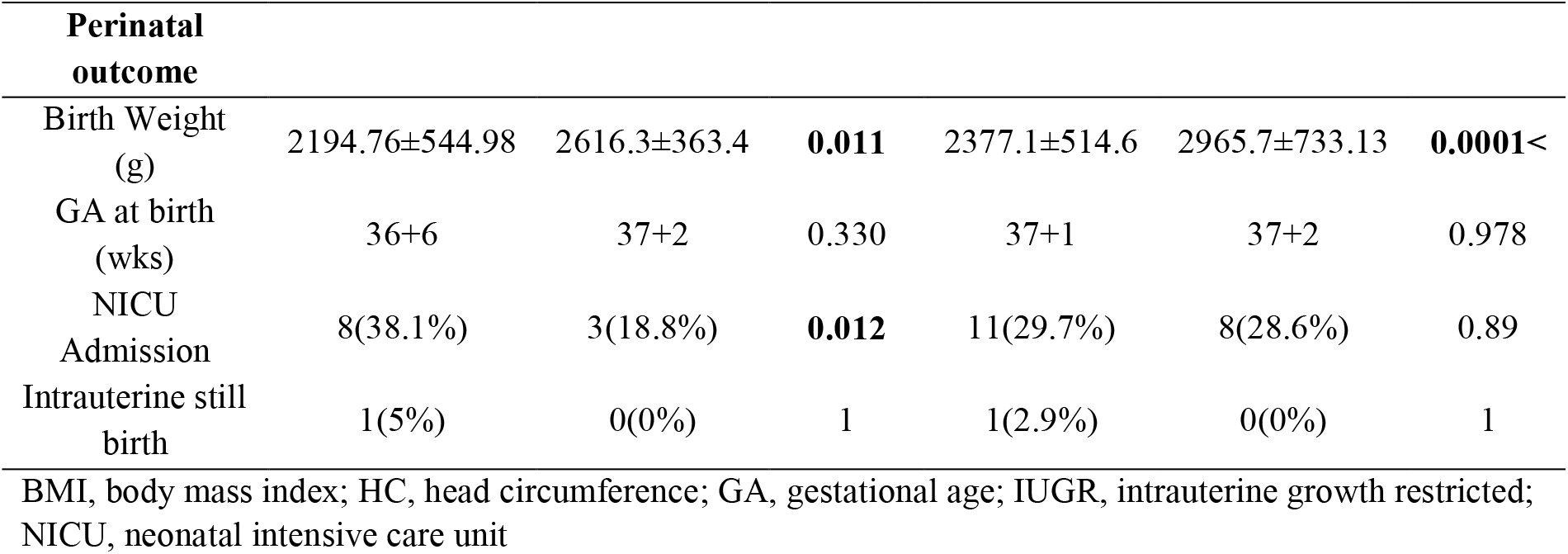
Maternal and fetal characteristics and perinatal outcome of the study groups.

### MRI examination

Brain white and grey matter signal and morphology were normal in all fetuses and no obvious hypoxic ischemic events as hemorrhagic changes were detected. Corrected cortical thickness in the insula and temporal lobes were significantly thinner in the IUGR group in comparison with the control group (0.034 vs 0.043 and 0.036 vs 0.047 respectively, P value <0.05) (Figure. 3). But there was no significant difference in frontal and occipital lobes cortical thickness (Table. 2). Insular, frontal and occipital corrected cortical thickness was thinner in subtype A than subtype B but the difference was not significant (Table. 3).

**Figure. 3.**
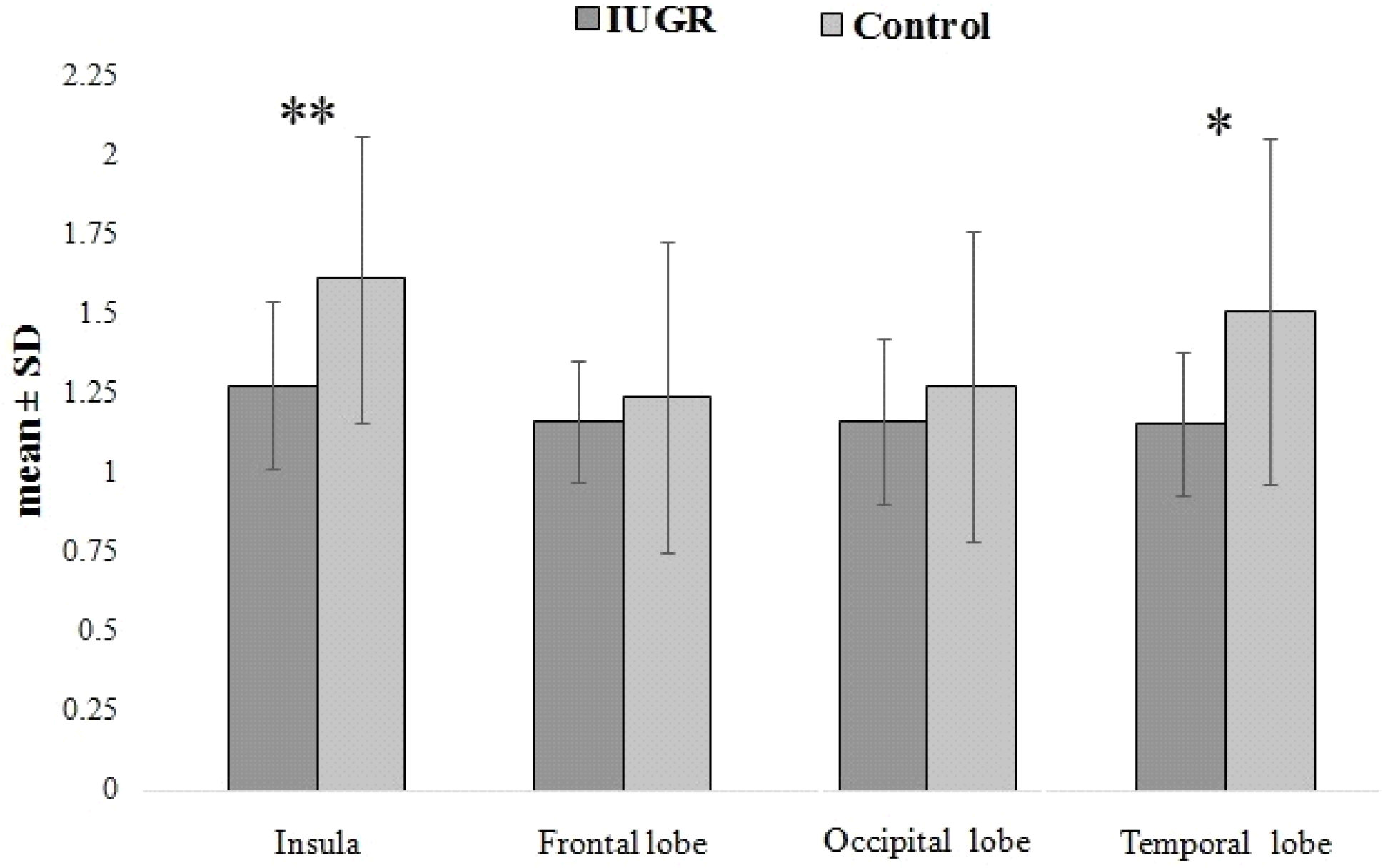
The polynomial contrast analysis figure demonstrates the difference of corrected cortical thickness in different brain regions between the IUGR group and Controls. The bars show mean±SD for each group. Asterisks demonstrate P<0.05

**Table 2:** Corrected brain area and corrected cortical thickness in IUGR group and controls.

|  | IUGR<br>n=42 | Control group<br>n=28 | P Value |
| --- | --- | --- | --- |
| **Cortical thickness(mm)** |  |  |  |
| Insula | 0.034±0.007 | 0.043±0.012 | **0.001** |
| Frontal lobe | 0.031±0.005 | 0.033±0.013 | 1 |
| Occipital lobe | 0.031±0.007 | 0.034±0.013 | 0.250 |
| Temporal lobe | 0.036±0.007 | 0.047±0.027 | **0.010** |
| **Area(cm2)** |  |  |  |
| Frontal lobe | 0.214±0.028 | 0.209±0.029 | 0.374 |
| Temporal lobe | 0.086±0.024 | 0.086±0.022 | 0.611 |
| Occipital lobe | 0.105±0.024 | 0.100±0.016 | 0.274 |
| Hippocampus | 0.017±0.008 | 0.0125±0.003 | **0.001>** |
| Cerebellum | 0.147±0.021 | 0.130±0.019 | **0.001** |
| Midbrain | 0.043±0.015 | 0.043±0.007 | 0.983 |
| Pons | 0.031±0.014 | 0.028±0.003 | 1 |
| Whole brain area | 0.806±0.170 | 0.812±0.027 | **0.028** |

**Table 3:** Difference of corrected brain area and corrected cortical thickness in group A IUGR and group B IUGR.

|  | Group A IUGR<br>n=24 | Group B IUGR<br>n=18 | P Value |
| --- | --- | --- | --- |
| **Cortical thickness(mm)** |  |  |  |
| Insula | 0.032±0.005 | 0.036±0.009 | 0.104 |
| Frontal lobe | 0.029±0.003 | 0.032±0.007 | 0.104 |
| Occipital lobe | 0.030±0.006 | 0.032±0.007 | 0.562 |
| Temporal lobe | 0.037±0.006 | 0.036±0.007 | 0.415 |
| **Area(cm2)** |  |  |  |
| Frontal lobe | 0.208±0.029 | 0.222±0.024 | 0.169 |
| Temporal lobe | 0.085±0.015 | 0.088±0.034 | 0.668 |
| Occipital lobe | 0.105±0.021 | 0.105±0.028 | 0.627 |
| Hippocampus | 0.015±0.003 | 0.019±0.012 | 0.334 |
| Cerebellum | 0.146±0.022 | 0.148±0.019 | 0.791 |
| Midbrain | 0.045±0.019 | 0.041±0.007 | 0.368 |
| Pons | 0.028±0.011 | 0.034±0.017 | 0.44 |
| Whole brain area | 0.785±0.220 | 0.831±0.049 | 0.566 |

IUGR fetuses have significantly smaller corrected WBA (0.806 vs 0.812, P value 0.028). The corrected area of the cerebellum and hippocampus were smaller in normally-grown fetuses (0.130 vs 0.147, 0.0125vs 0.017, respectively, P value of < 0.05) (Figure 4), but other corrected areas of the brain were not significantly different between IUGR and control group (Table. 2). The corrected area of frontal, temporal, cerebellum and hippocampus in subtype A were smaller than subtype B but the difference was not significant (Table. 3).

**Figure. 4.**
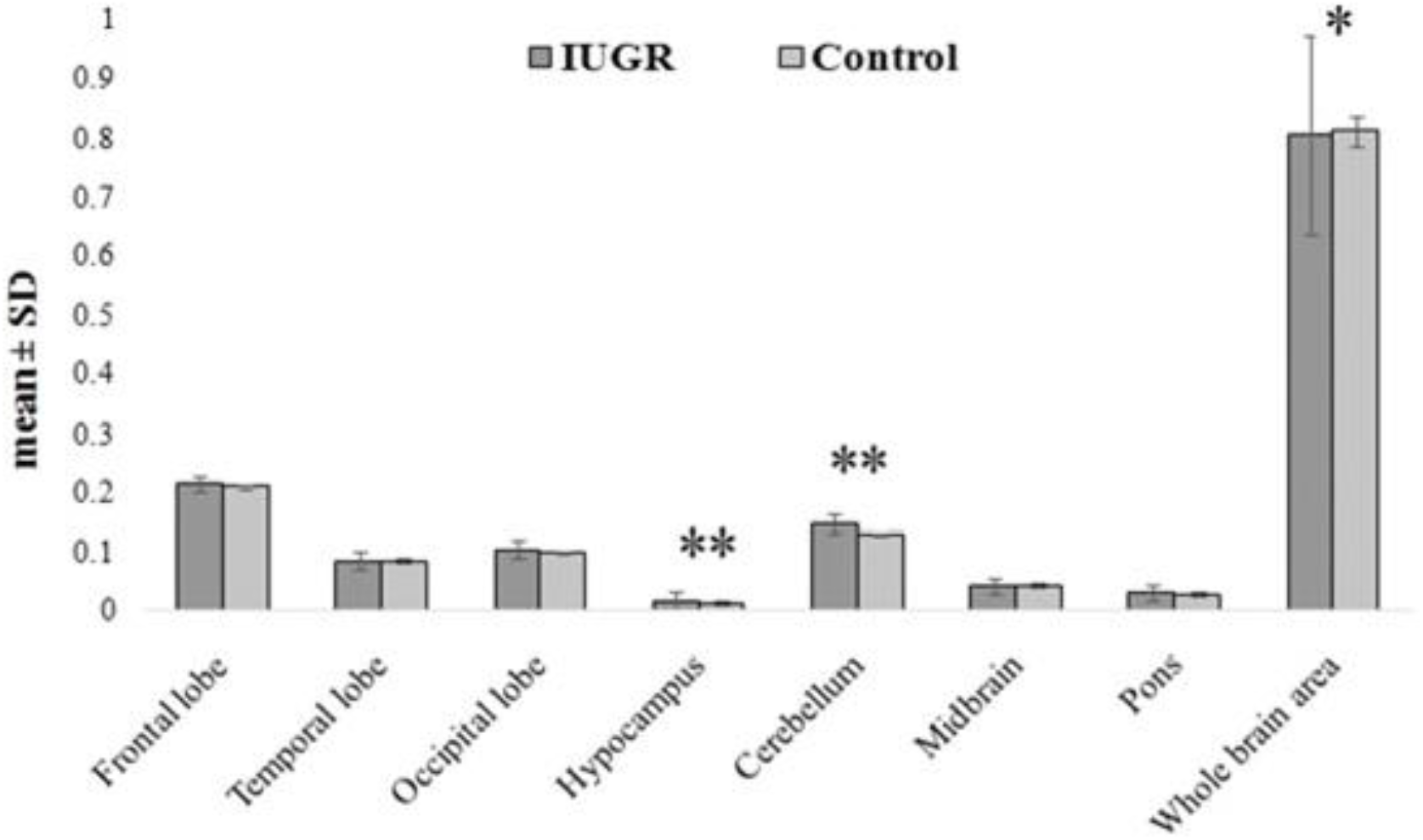
The polynomial contrast analysis figure demonstrates the difference between corrected brain areas in different lobes between the IUGR group and controls. The bars show mean±SD for each group. Asterisks demonstrate P<0.05

## Discussion

Our study has demonstrated that corrected cortical thickness in the insula and temporal lobes were significantly thinner in IUGR fetuses compared to the control group. Also, the whole brain area was significantly smaller in the IUGR group. Interestingly, the corrected areas of the cerebellum and hippocampus were smaller in controls.

Several studies suggest that IUGR due to placental insufficiency is associated with specific structural and functional changes in cortical development of the brain (9,5). IUGR neonates have a delay in cortical development at birth and also have brain cortical thickness and volume reduction (3, 20).

The insula is a main part of the limbic system and has the main role in cognition and awareness (20). In our study, the corrected cortical thickness of insula is thinner in IUGR fetuses compared to the control group. Our results are compatible with Egana-Ugrinovic et al. study in which it was shown that IUGR fetuses have reduced insular cortical thickness compared to controls. The possible mechanism is that this area is vulnerable to long term hypoxia (8).

Moreover, our IUGR group had significantly thinner corrected temporal cortical thickness. Our results are in line with another study that showed bilateral reduced gray matter in the temporal lobe in IUGR fetuses compared with the controls (14). More researches need to be conducted to obtain a better interpretation of the effect of growth restriction on temporal cortical thickness.

Our results show that the whole brain area in the IUGR group was significantly lower than controls and our findings are compatible with previous studies (5,9,15,22). These differences may be due to the effect of placental insufficiency on DNA synthesis, reduction in brain cell size and number, synaptogenesis and total brain weight (23,24). However, in contrast to the abovementioned studies, there was only one research that found no significant difference in whole brain volume between the IUGR and normal fetuses (25).

It is worth to note that in this research, the corrected area of the cerebellum was larger in the IUGR group compared to controls. We assessed the corrected area of the cerebellum by dividing the cerebellar area by whole brain area and as mentioned whole brain area in IUGR groups was smaller. This finding may show that the cerebellum is less affected by IUGR compared with other brain regions. Our results are approximately in line with a study by Sanz-Cortes et al., who assessed corrected cerebellar volume by dividing cerebellar volume to whole brain volume, they showed significantly larger cerebellar ratio in IUGR fetuses (26,27). In addition, Bruno et al showed that there was no significant difference in cerebellar volume between IUGR and control groups (27). Research using ultrasound also showed preservation of trans cerebellar diameter in IUGR fetuses (28). All the above-mentioned studies emphasize the preservation of cerebellar size in IUGR fetuses. However, research by Polat et al. found decreased cerebellar /supratentorial volume ratio in IUGR fetuses (29) and another study by Anderscavage et al. demonstrated smaller cerebellar volume in IUGR fetuses (30). Due to variable results, further studies are needed to evaluate the growth restriction effect on fetal cerebellum.

Hippocampus is an important grey matter structure and is sensitive to placental insufficiency, hypoxia, lower nutritional supply and maternal stress (31,32,33). An animal study showed adverse molecular and cellular effects of IUGR on hippocampus structures (34). Interestingly, in our study IUGR fetuses had significantly larger corrected hippocampus area and similar to the cerebellar area, this region may be less affected by IUGR. Padilla et al studied a sample of IUGR and AGA neonatal groups, they found no difference in hippocampus volume between these groups (15). On the contrary, Gregory et al. suggested that total hippocampal volume is affected by IUGR, they reported significantly smaller hippocampus volume in IUGR fetuses compared with controls (7).

Our study showed no statically significant correlation between frontal and occipital corrected areas among IUGR and control groups. No study was found to evaluate different brain lobe areas, but our study is in line with the research of padilla et al. that demonstrated similar frontal and occipital relative volumes between IUGR fetuses and controls (15). Some studies in older age showed smaller frontal lobe volumes in IUGR group and the difference in relative frontal volume may no longer observable up to 1 year old (15). On the other hand, a study using sonographic biometry showed decreased frontal dimension in IUGR neonates compared to controls (35). It has been demonstrated that the frontal lobe may have delayed growth pattern in IUGR (34). Considering these findings, the preservation of the frontal lobe area in IUGR fetuses of our study may be due to developmental process of frontal lobe and the lower gestational age of evaluation.

This study had some limitations. First, the sample size was relatively small, especially for comparing the faint differences between IUGR subtypes. Second, we manually traced all brain areas and it could be subject to inter or intra-reader variability. Third, the number of IUGR fetuses and control group were not similar. The most non-IUGR cases referred for fetal MRI at lower gestational age opposite to the IUGR cases. Therefore, for matching the gestational age of two groups we had limitation in including of control fetuses. And at the end, because our control group had been selected from fetuses with other pathologies, the post-natal outcome was not different between IUGR and control groups.

## Conclusion

Our study shows that IUGR has selective effects on brain morphometrics. IUGR fetuses have a thinner cortical thickness in insular and temporal lobes and smaller whole brain areas. Furthermore, cerebellum and hippocampus were less affected by growth restriction. Further detailed investigations in the future will be needed to evaluate morphometrics effects of IUGR on the insula, cerebellum, hippocampus, and temporal lobe and its association with neurological outcome.

## Data Availability

THE DATA ARE AVAILABLE

## Acknowledgments

This study was financially supported by the Radiology research center, Imam Khomeini Hospital, Tehran, Iran (grant number: 98-01-98-41442)

